# Correction in Active Cases Data of COVID-19 for the US States by Analytical study

**DOI:** 10.1101/2020.09.02.20186833

**Authors:** Ravi Solanki, Anubhav Varshney, Raveesh Gourishetty, Saniya Minase, Namitha Sivadas, Ashutosh Mahajan

## Abstract

The reported recovered cases for many US states are low which could be due to difficulties in keeping track of recoveries and resulted in higher numbers for the reported active cases than the actual numbers on the ground. These incorrect numbers can lead to misleading inferences. In this work, based on the typical range of recovery rate of COVID-19, we estimate the active data from the total cases and death cases and bring out a correction for the data for all the US states reported on worldometer.

## BACKGROUND

The availability of accurate data of an epidemic is important as the data provides key insights on the disease spread and enable the authorities to take a decision on control measures. Worldometer is one of the very popular sources of the global COVID-19 data and it is also trusted and used by many government bodies and agencies [1]. The available data for the COVID-19 cases can be used for the prediction and analysis of hospitalization and meeting the demands of health care facilities and setting up the critical care systems for the patients [2] [3]. The active cases represent the number of infected people whether symptomatic or asymptomatic detected through self-reporting or testing. This number is important for Public Health authorities to estimate the current status of the disease spread and can be calculated by subtracting death and recovered cases from the total confirmed cases.

## METHOD

A compartmental predictive mathematical model SIPHERD for COVID-19 disease dynamics is described in [4], [5] where recovery rate is a model parameter and is fixed by optimizing the model with the actual data. The data for the total, death, and active cases for 168 days from March 4, 2020, is taken from Worldometer [1] and found close to total cases and death data from [6]. After running the SIPHERD model for the USA, as reported in [4], the recovery rate of the active category was found to be 0.015 (corresponding to 66 days of mean recovery time) which is very low compared to other countries like Germany and India [4] [5] where it is found 0.065. The low recovery rate in the USA may be attributed to either incorrect reporting of the Active cases [7] or the testing of serious cases only and longer recovery time in hospitals compared to quarantined with mild symptoms. Secondly, keeping the record of recoveries is difficult because some of the infected people are asked to quarantine while only critical patients are hospitalized. Sometimes the reporting of those recoveries is not accurate or incomplete. This has led to inconsistent data for active cases.

The number of mild cases is reported to be 81% in a Chinese study [8]. Covid-19 data reported from 49 states, the District of Columbia, and three U.S. territories to CDC from February 12–March 16 shows that 20.7 reported cases were severe and were hospitalized [9]. COVID-NET regions show this number to be 21.4 % till April 4 [10], [11] and IHME data March 5- April 4 shows that to be 20.3 % [12] [13]. According to WHO recovery time for mild cases is of 2 weeks and for severe cases that recovery time of 3-6 weeks. [14]. Considering 80% mild cases, the recovery rate cannot be as low as appears from the data for the states listed in column 2 in Table 1.

The correct estimation of the active cases can be done by subtracting the death and recovery cases, with appropriate recovery rate, from the total cases. The woldometer data for total cases and death cases are assumed to be true as the test positive results and count of diseased cases are done more stringently as compared to recovery counting. The active cases can be obtained by solving the following differential equation,

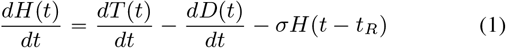

where, *H, T*, and *D* are the active, total, and death cases. The recovery from the infected category is defined by the two parameters: delay in recovery *t_R_* and the recovery rate *σ*. As these values are dependent upon the immune system of the community and the hospital facilities, it should not vary much within the USA. We have taken *t_R_* as 10 days and *σ* as 0.048 (21 days of mean recovery time considering both mild and severe cases).

## RESULTS AND DISCUSSION

The above delay differential equation is solved for all the states of the USA and we found three groups among the states according to the accuracy of the data. The active cases reported on worldometer [1] for a few states show excellent agreement with our estimation of active cases. One example state for this group is Texas as seen in Fig. 1(a). There are few states in the second group which are largely not matching with analytical estimation indicating that reported active data is inaccurate. These states are listed in column 2 in Table 1 and one representative state is Virginia as seen in Fig. 1(b), where the currently active cases are reported to be 97560 which should have been just around 21664 according to our calculation. Interestingly, in the last group, there are some states for which the reported active cases do not follow the estimated active cases for the initial days, however, the reported data of the active cases take a sudden fall and follow our estimation as seen for Indiana in Fig. 1(c). This clearly indicates that the active cases data corrections is done after some days and also validates our analytical approach. In Fig. 1, the reported total and active cases with the estimated active cases for one of the states in all the three groups are shown, the figures for remaining states are given in supplementary material.

**Fig. 1:**
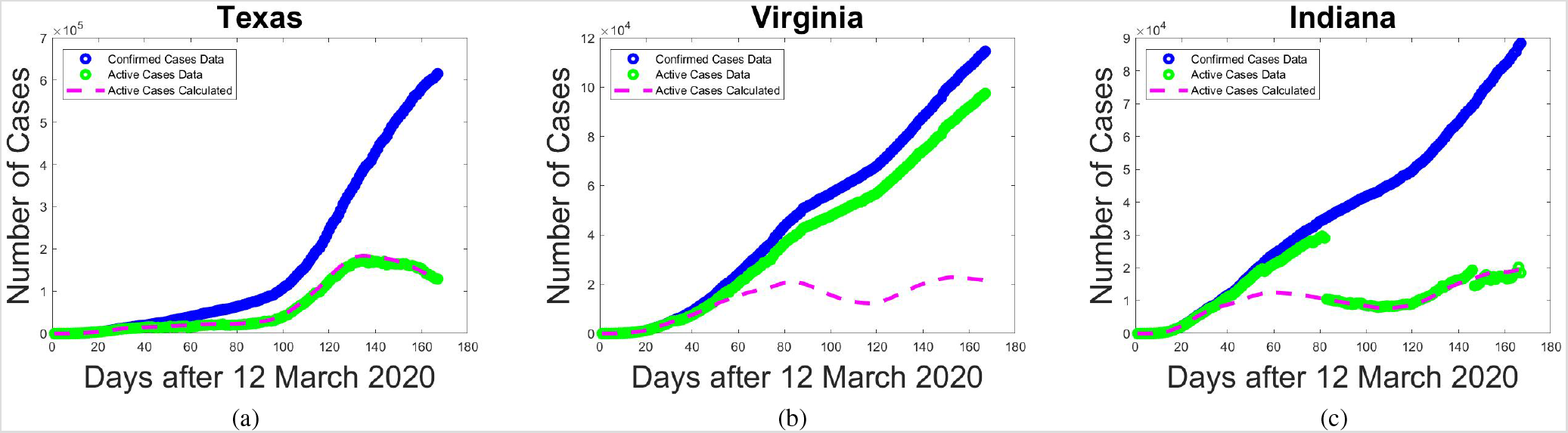
(a) Texas representing states in which active data is corrected reported. (b) Virginia represents the second group where data is largely incorrect and (c) Indiana from the third group where data has been corrected recently and closely match with analytical estimation.

**TABLE 1:**
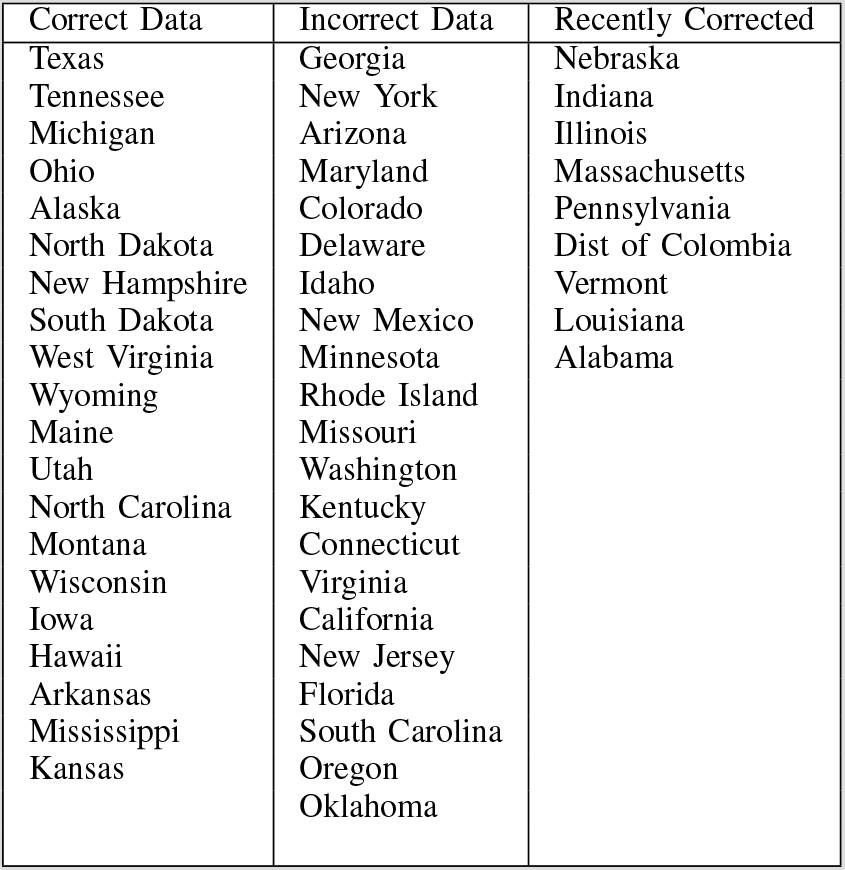
The US States Active Cases Data Status

| Correct Data | Incorrect Data | Recently Corrected |
| --- | --- | --- |
| Texas | Georgia | Nebraska |
| Tennessee | New York | Indiana |
| Michigan | Arizona | Illinois |
| Ohio | Maryland | Massachusetts |
| Alaska | Colorado | Pennsylvania |
| North Dakota | Delaware | Dist of Colombia |
| New Hampshire | Idaho | Vermont |
| South Dakota | New Mexico | Louisiana |
| West Virginia | Minnesota | Alabama |
| Wyoming | Rhode Island |  |
| Maine | Missouri |  |
| Utah | Washington |  |
| North Carolina | Kentucky |  |
| Montana | Connecticut |  |
| Wisconsin | Virginia |  |
| Iowa | California |  |
| Hawaii | New Jersey |  |
| Arkansas | Florida |  |
| Mississippi | South Carolina |  |
| Kansas | Oregon |  |
|  | Oklahoma |  |

The recovery rate in individual states may vary to an extent of ±10% depending on the variation in the number of tests performed, fraction of mild and severe cases. However, we have taken a uniform value of recovery rate as 0.048 for all the US states.

## CONCLUSIONS

The reported active cases for a few states is consistent with the total detected cases and death cases for a recovery rate parameter value 0.048. For a few states, the data has been corrected recently. However, for 21 states, the active cases data is still largely incorrect. We generate the corrected active case data for all states, report in the supplementary material, and also keep it available on Github.

## Data Availability

Total, active and death cases data for COVID-19 is downloaded from the publicly available data sources.
https://www.worldometers.info/coronavirus/country/us/

https://github.com/ravisolankigithub/covid-activecases-usa.git

## A. Data Availability

The data for the corrected active case for all the US States can be downloaded with the github link below.

https://github.com/ravisolankigithub/covid-activecases-usa.git

## SUPPLEMENTARY MATERIAL

**Fig. S1:**
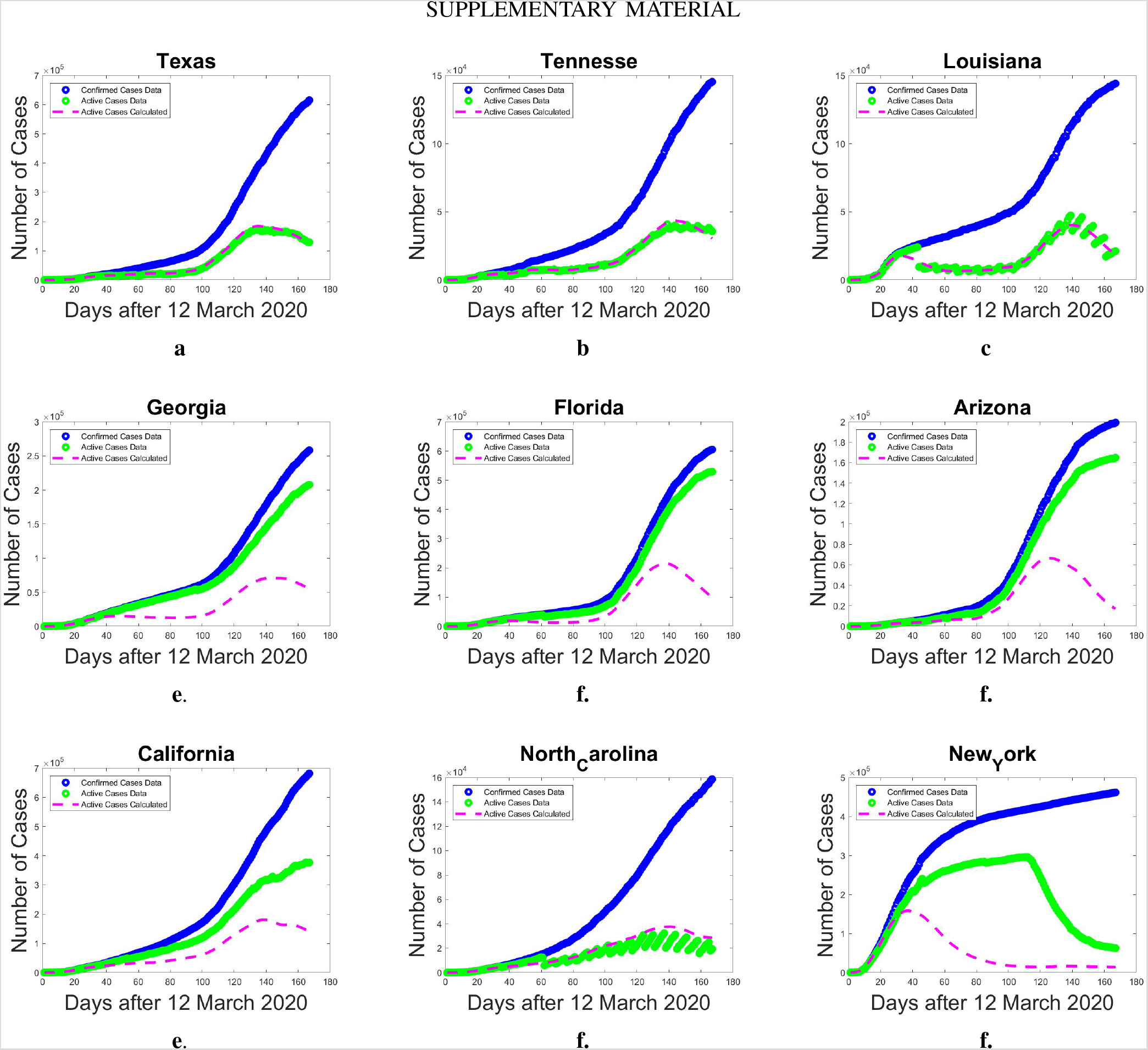
Reported total cases, active Cases data and analytically estimated active cases for various US states

**Fig. S2:**
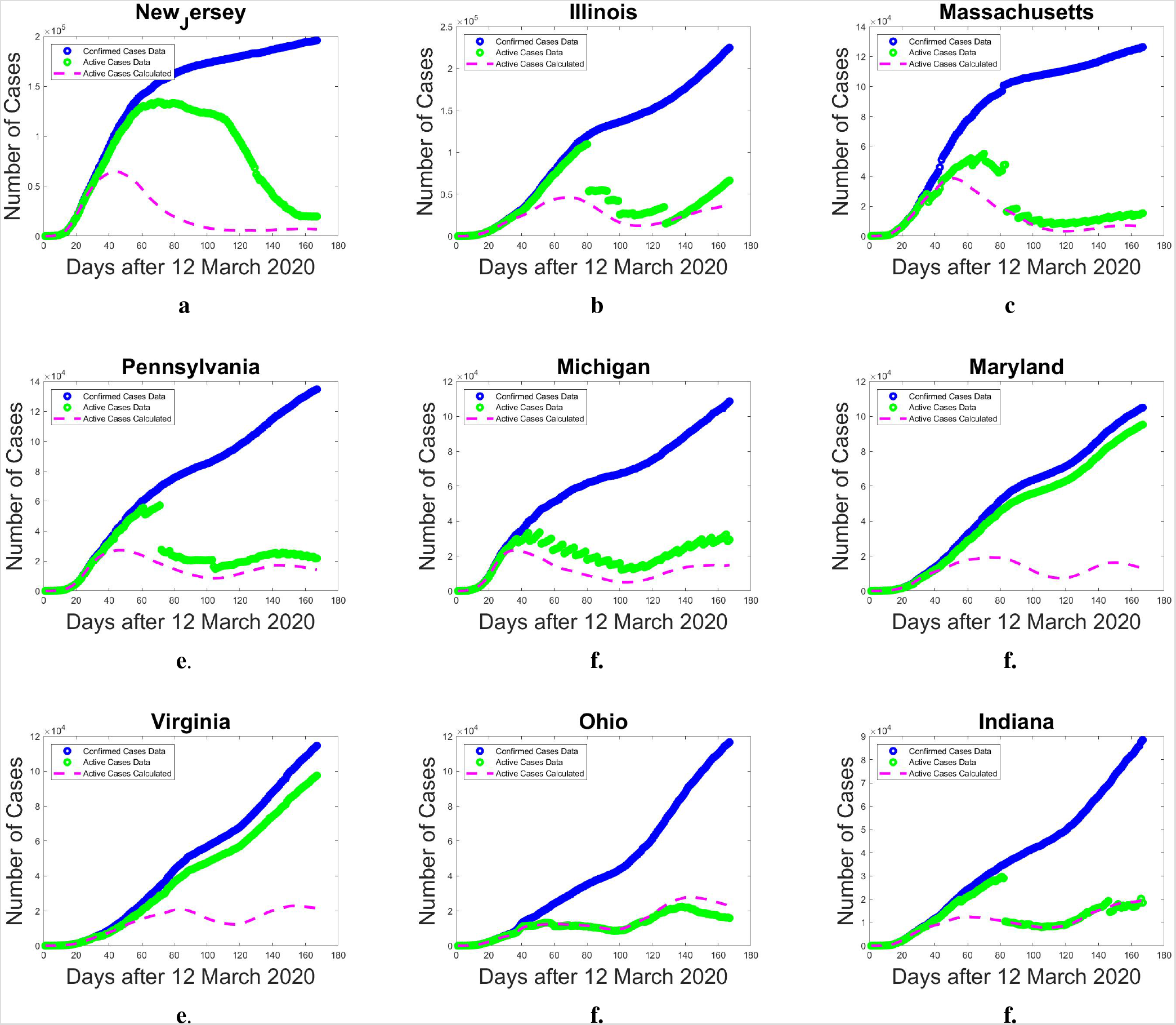
Reported total cases, active Cases data and analytically estimated active cases for various US states

**Fig. S3:**
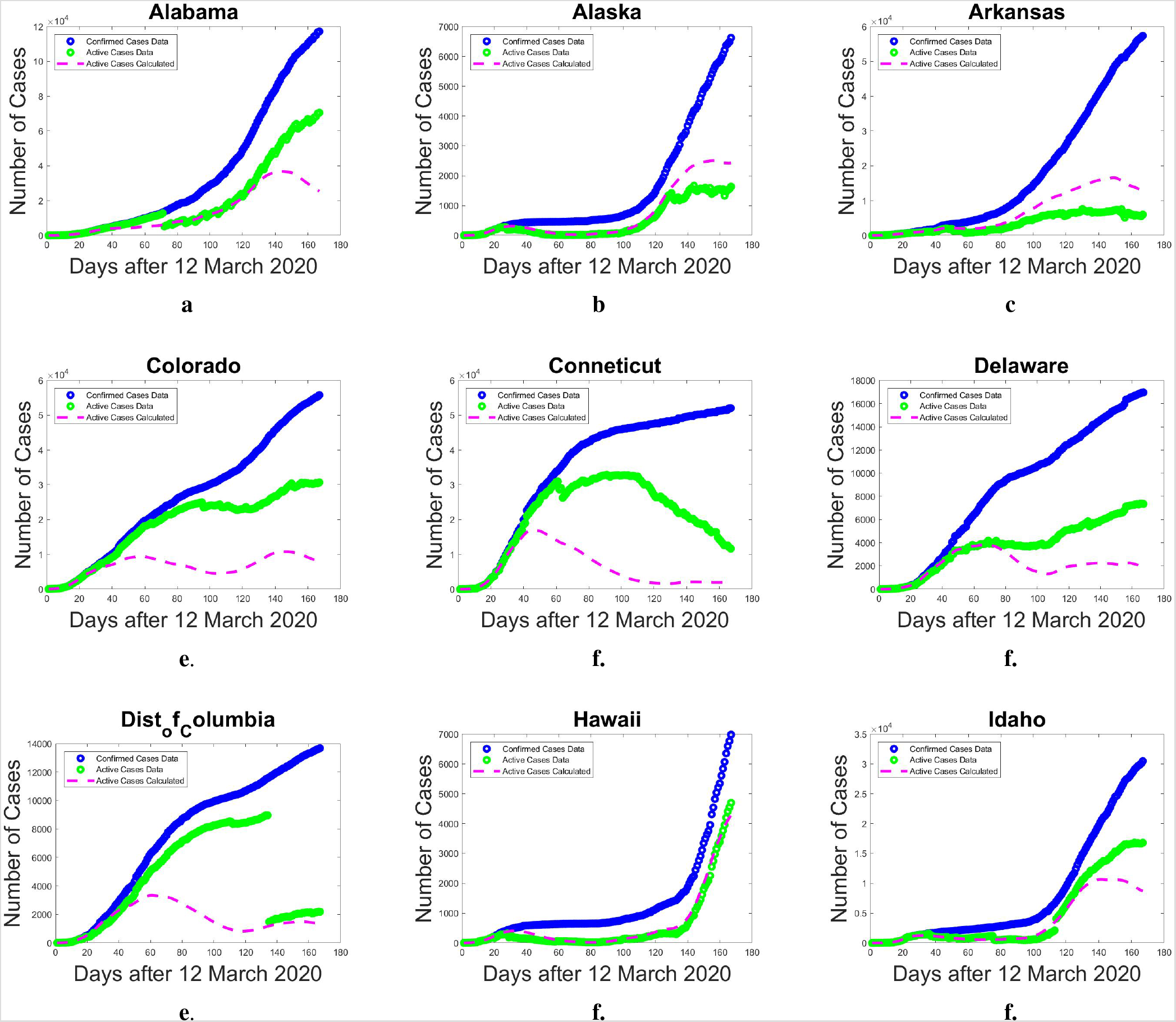
Reported total cases, active Cases data and analytically estimated active cases for various US states

**Fig. S4:**
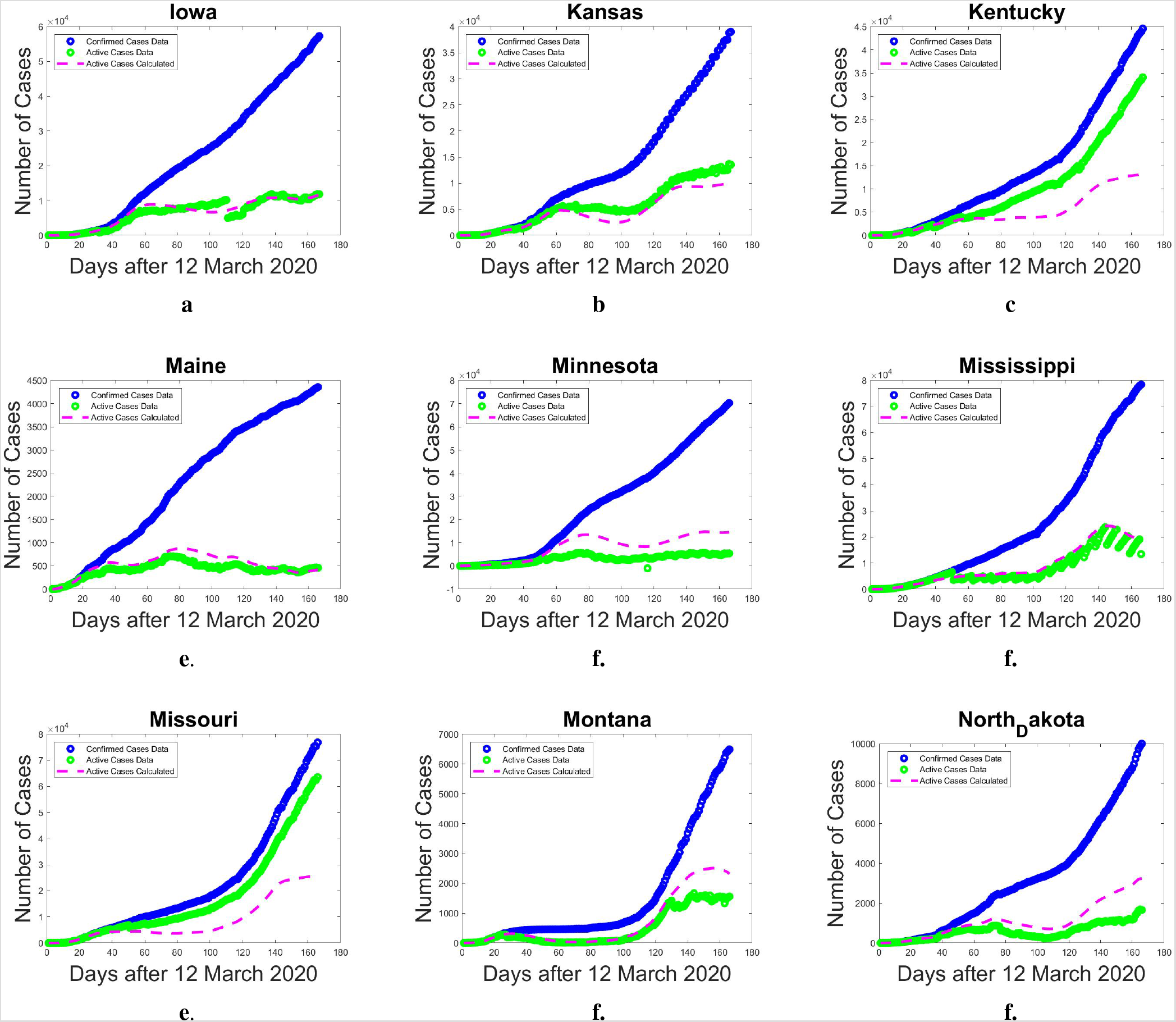
Reported total cases, active Cases data and analytically estimated active cases for various US states

**Fig. S5:**
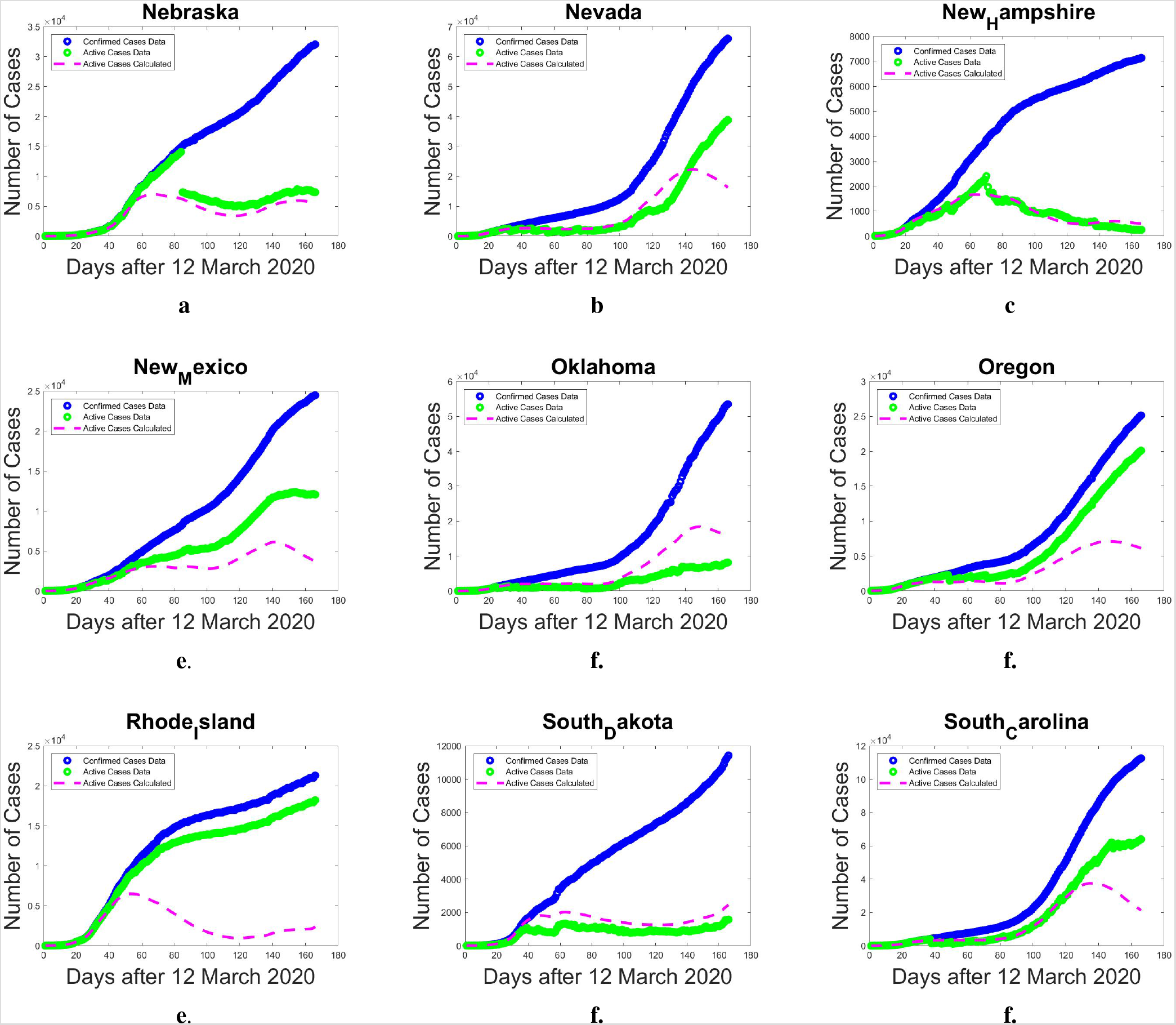
Reported total cases, active Cases data and analytically estimated active cases for various US states

**Fig. S6:**
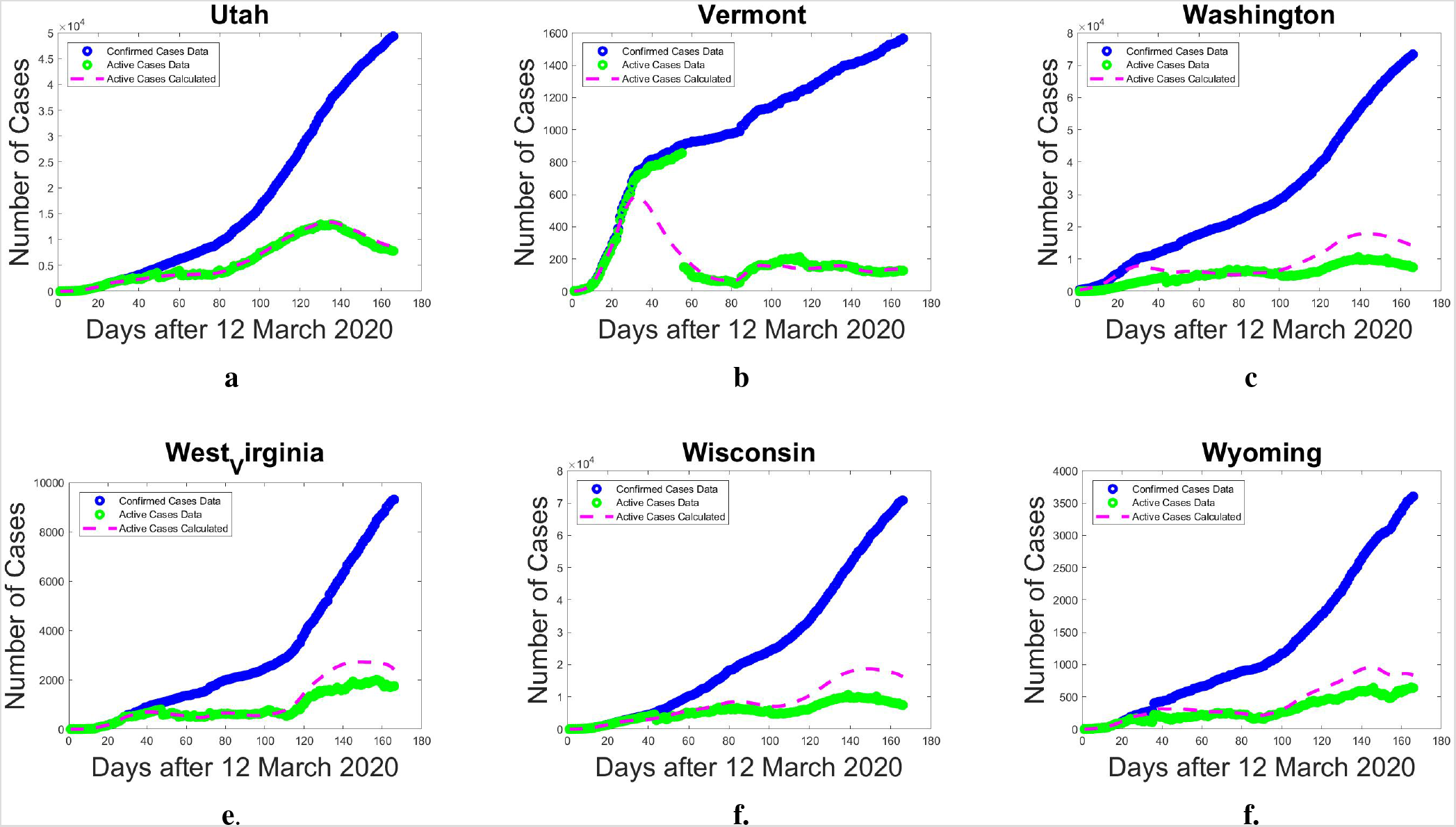
Reported total cases, active Cases data and analytically estimated active cases for various US states

